# Comparing the Performance of HIV Rapid Diagnostic Tests used in Zambia - A Systematic Clinical Data Review

**DOI:** 10.1101/2021.12.20.21267838

**Authors:** L. Mukuka, A. Theo, M. Zambwe, P. J. Chipimo

## Abstract

**Objective:** To investigate the performance of the HIV RDTs used in Zambia.

**Method:** 2,564 participants aged between 15 and 95 years from two sites in Lusaka province years were tested on OraQuick ADVANCE, Abbot Determine™, and then confirmed on Uni-Gold™ Recombigen®. The data from the participants were analyzed using SPSS version 25.0.

**Results:** The 3 RDTs when compared to the 4^th^ generation Abbot Architect results had the following results: OraQuick *ADVANCE*®, Alere Determine and Uni-Gold Ultra, at 95% CI had Sensitivities of: 91.8%, 93.3% and 92.5% respectively. The specificities of OraQuick *ADVANCE*® and Uni-Gold were the same (100.0%; 95% CI: 98.8 -100.0) but slightly different from Alere Determine (99.8%). Positive predictive values at 95% CI were 100% for OraQuick *ADVANCE*® and Uni-Gold and 98.4% for Alere Determine. Negative predictive values (at 95% CIs) were 99.1, 99.2 and 99.1 for OraQuick *ADVANCE*®, Alere Determine, and Uni-Gold Ultra respectively. The results showed that these RDTs could only detect 12 out of every 13 HIV positive results.

**Conclusion:** Third generation RDTs are not effective in detecting acute positive cases. Fourth generation Rapid Tests are required to capture the positive cases being missed out.

## INTRODUCTION

According to WHO global HIV summary report, the HIV epidemic is still raging thus emphasizing the need for a sustained response in order to curb the HIV epidemic.^1^ Despite all the strategies put in place to help end the HIV/AIDS epidemic, there still remains a substantial number of people with HIV who are unaware of their infection and are still infecting others.^2^ The only way to determine a person’s HIV status is to have an HIV test. Globally about 20% of HIV infections are due to transmissions from recently infected individuals.^3^ The World Health Organization recommends two sequential rapid diagnostic tests (RDTs) for HIV diagnosis, however third generation RDTs detect only HIV antibodies and may miss up to 75% of early acute HIV infections.^4^

There is considerable interest in revising testing guidelines to more accurately reflect new technology and to identify associated challenges.^5^ According to Marks et al the estimated 25% of individuals unaware of their HIV infection are responsible for 54% of new infections. Point of care (POC) options for the diagnosis of acute HIV are currently limited to third-generation RDTs.^6^ An addition of a fourth generation HIV RDT to testing algorithms would help in reducing the number of acute infections being missed by third generation RDTs that only detect antibodies. Identification of acute HIV infection requires detection of HIV nucleic acids or p24 antigens which is made possible with 4^th^ generation laboratory-based assays that detect HIV-1 p24 antigen as well as antibodies to HIV-1/2.^7, 8, 9^ Many studies have shown that fourth-generation RDTs can still detect a good number of acute HIV infections ^11^ compared to third-generation RDTs.

The reported sensitivity of a test also depends on the sensitivity of the comparator or reference standard.^12^ Manufacturers of four commonly studied HIV rapid tests quote sensitivities of 99.3 to 100%, but all were compared with earlier Enzyme Immuno-Assays (EIA) that are less sensitive than the now widely available fourth-generation EIAs.^12^ Fourth-generation EIAs can detect HIV p24 antigen as early as 2 weeks post-infection, as well as being able to detect the antibodies that appear later.^12^ These EIAs are recommended as first-line screening tests in the 2014 Centers for Disease Control and Prevention (CDC) HIV laboratory testing algorithm.

Although the proportion of acute infections can be expected to vary across different populations, there are no studies examining the effect of this on the performance of HIV rapid tests.^13^ Health services that have the option of providing either HIV rapid tests or the more sensitive laboratory assays therefore need to know whether there will be a significant loss of sensitivity if they use HIV rapid tests in their clinical population.^13^

Zambia has an adult HIV prevalence of 11.1%. The national RDT algorithm in Zambia consists of a screening test (Determine® HIV 1/2) followed by confirmation of reactive specimens with a second rapid test (Uni-gold HIV 1/2). ^14^ This study set out to investigate the performance of the HIV RDTs used in Zambia.

**Figure 1:**
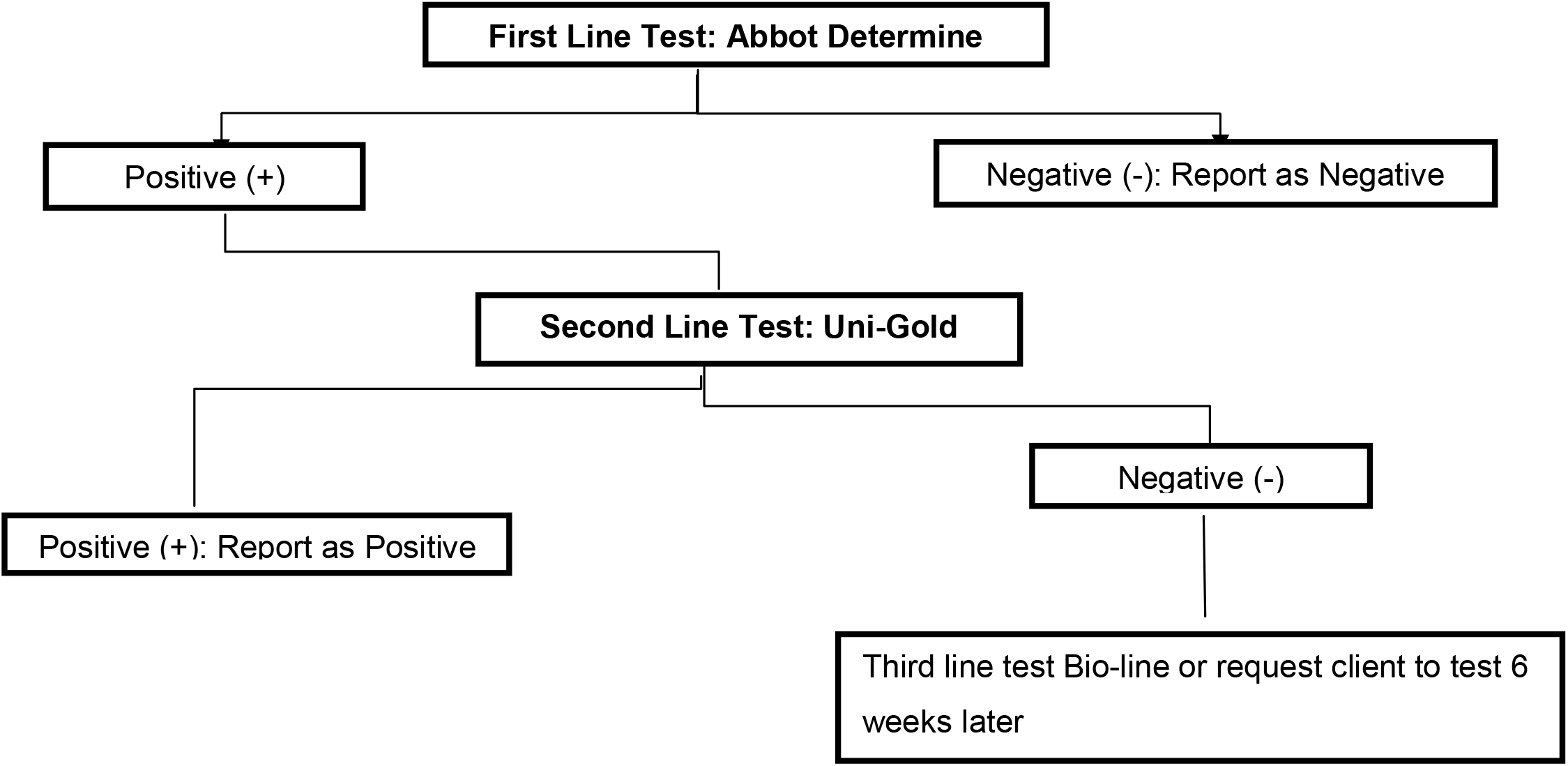
Zambia National algorithm for HIV testing using rapid tests.

## METHODS

This systematic review used quantitative methods by measuring statistical analysis of different variables of the five different HIV tests by using probability methods. The comparison between these diagnostic tests forms the foundation of this comparative evaluation of HIV testing techniques. Serologic test results were divided into Reactive and Non-Reactive findings, as recommend by the manufacturers of the diagnostic tests. This study analyzed data collected from the STAR clinical performance study.^15^The analysis is solely dependent on the secondary data from the STAR study. Serologic methodologies for the 5 test methods were evaluated for HIV. The primary purpose of this investigation was to complete a comparative evaluation of 3 HIV rapid diagnostic tests and 2 fourth generation tests.

The specimen was collected by trained personnel following participant informed consent and complete assessment. Samples analyzed included oral mucosa transudate using the OraQuick® HIV Self-Test, fingerprick blood was collected and tested on Alere Determine™ HIV1/2, if positive on Determine it was then confirmed on Unigold™HIV1/2 (Trinity Biotech) test following the Zambian national testing algorithm. The results were provided to participants, and data stored in electronic devices, all participants with reactive test results were referred for HIV care and treatment at local health facility.^15^

In addition to the three rapid tests performed at Point of Care, 10 mls whole blood was collected from each participant by trained personnel and sent to the Laboratory for further testing on fourth generation assays. Specimens were processed by standardized methods and tested following the manufacturer’s instructions for all the diagnostic test methods. The 10mls blood samples sent to the Laboratory were double-span and plasma was separated and tested on the Abbot Architect HIV Ag/Ab Combo fourth generation instrument as the gold standard for the rapid tests.^15^ If result was reactive, it was then run on manual ELISA Genscreen™ ULTRA HIV Ag-Ab to confirm positivity.

Specificity and Sensitivity of the five different HIV testing strategies were measured by calculating the percentage of positive results yielded by these tests among all samples that tested positive for HIV based on detection of HIV positive results of the Abbot Architect test.^16^ Rapid test positivity was calculated by comparing the characteristics of HIV-positive and HIV-negative results. The Results were reported in accordance with the Standards for Reporting of Diagnostic Accuracy: Sensitivity, specificity, positive and negative predictive values at 95% confidence intervals using SPSS version 25.0. The gold standard serologic tests for HIV are the enzyme immunoassay (EIA). The Determine HIV1/2, product sensitivity and specificity calculations were based on the comparison of test results with an indirect enzyme immunoassay the Abbot Architect and Genscreen HIV-1/2.

### Inclusion and exclusion criteria

The study only included samples of participants who gave informed consent to participate in the STAR study by Zambart^15^ and who were tested on all 5 of the HIV test types being investigated. Participants who did not give consent or did not give a third sample for Laboratory testing were excluded from the study.

### Statistical analysis

Data was analyzed using Microsoft Excel spreadsheet for broader thematic analysis and SPSS version 25.0. Exact confidence intervals (CIs) were calculated according to the binomial distribution. Variables were summarized as frequencies and percentages with significant level at 0.05 (95%CI). Sensitivity, specificity, positive and negative predictive values were determined using cross-tables and standard formulas of the third generation HIV RDTs, the Abbot fourth generation immunoassay was used as the gold standard. Data from the STAR-Study were entered into a Microsoft excel sheet, checked for duplication and error and then subsequently exported to SPSS version 25 for analysis.

## RESULTS

The study examined specimen of 2564 participants, which were tested on the 5 different HIV tests being investigated. The rapid diagnostic laboratory tests were evaluated based on a comparison test method. The investigation evaluated the test accuracy of five diagnostic assays for the identification of HIV based on sensitivity and specificity measures. Among the 2564 samples analyzed, 1043 (40.7%) were male participants and 1521 (59.3%) female participants. The 2564 samples were from participants of a wide range of ages from the age of 15 to as old as 95 years as show in table 3. All participants below 18 years agreed to participate with guided parental consent.

**Table 1:**
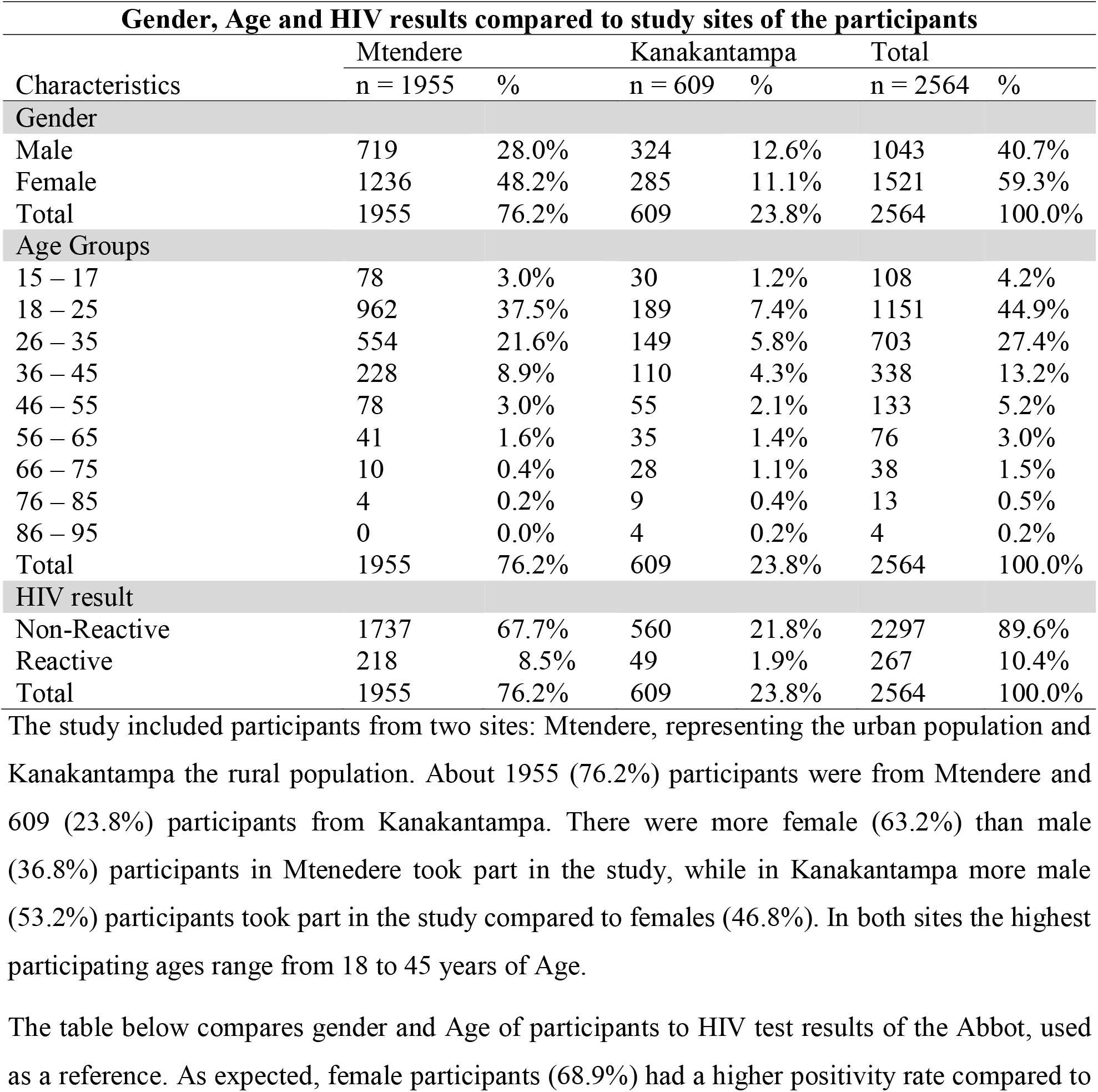
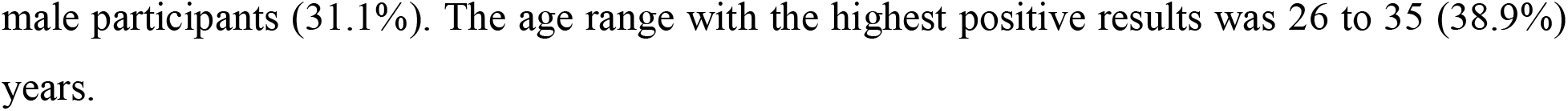
Gender, Age Groups and HIV Status on Abbot Architect of the study participants compared to Sites they lived.

**Table 2:**
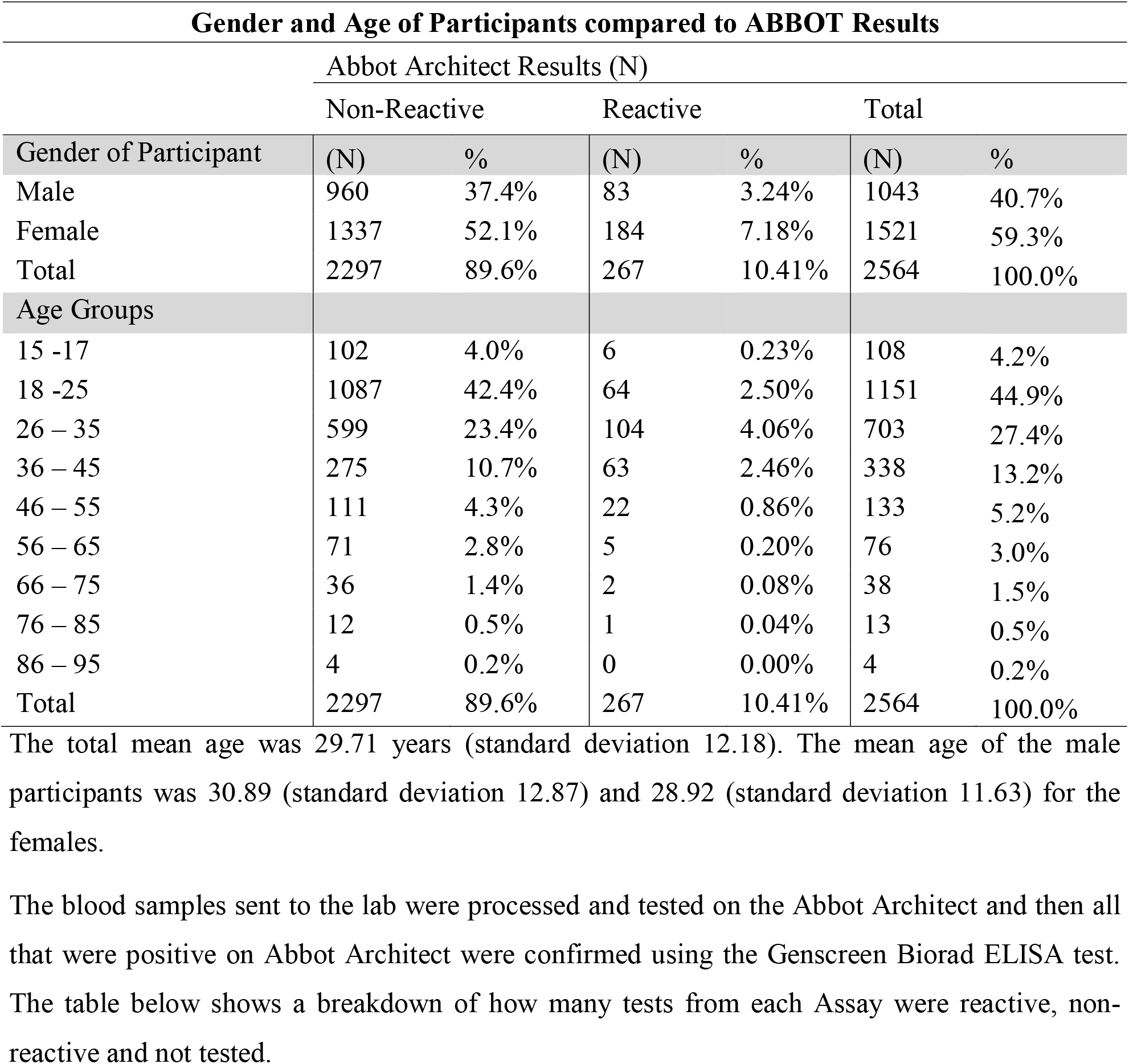
Gender and Age of Participants compared to the gold standard HIV test results.

| **Gender and Age of Participants compared to ABBOT Results** |  |  |  |  |  |  |
| --- | --- | --- | --- | --- | --- | --- |
| Abbot Architect Results (N) |  |  |  |  |  |  |
|  | Non-Reactive |  | Reactive |  | Total |  |
| Gender of Participant | (N) | % | (N) | % | (N) | % |
| Male | 960 | 37.4% | 83 | 3.24% | 1043 | 40.7% |
| Female | 1337 | 52.1% | 184 | 7.18% | 1521 | 59.3% |
| Total | 2297 | 89.6% | 267 | 10.41% | 2564 | 100.0% |
| Age Groups |  |  |  |  |  |  |
| 15 -17 | 102 | 4.0% | 6 | 0.23% | 108 | 4.2% |
| 18 -25 | 1087 | 42.4% | 64 | 2.50% | 1151 | 44.9% |
| 26 – 35 | 599 | 23.4% | 104 | 4.06% | 703 | 27.4% |
| 36 – 45 | 275 | 10.7% | 63 | 2.46% | 338 | 13.2% |
| 46 – 55 | 111 | 4.3% | 22 | 0.86% | 133 | 5.2% |
| 56 – 65 | 71 | 2.8% | 5 | 0.20% | 76 | 3.0% |
| 66 – 75 | 36 | 1.4% | 2 | 0.08% | 38 | 1.5% |
| 76 – 85 | 12 | 0.5% | 1 | 0.04% | 13 | 0.5% |
| 86 – 95 | 4 | 0.2% | 0 | 0.00% | 4 | 0.2% |
| Total | 2297 | 89.6% | 267 | 10.41% | 2564 | 100.0% |

The total mean age was 29.71 years (standard deviation 12.18). The mean age of the male participants was 30.89 (standard deviation 12.87) and 28.92 (standard deviation 11.63) for the females.

**Table 3:**
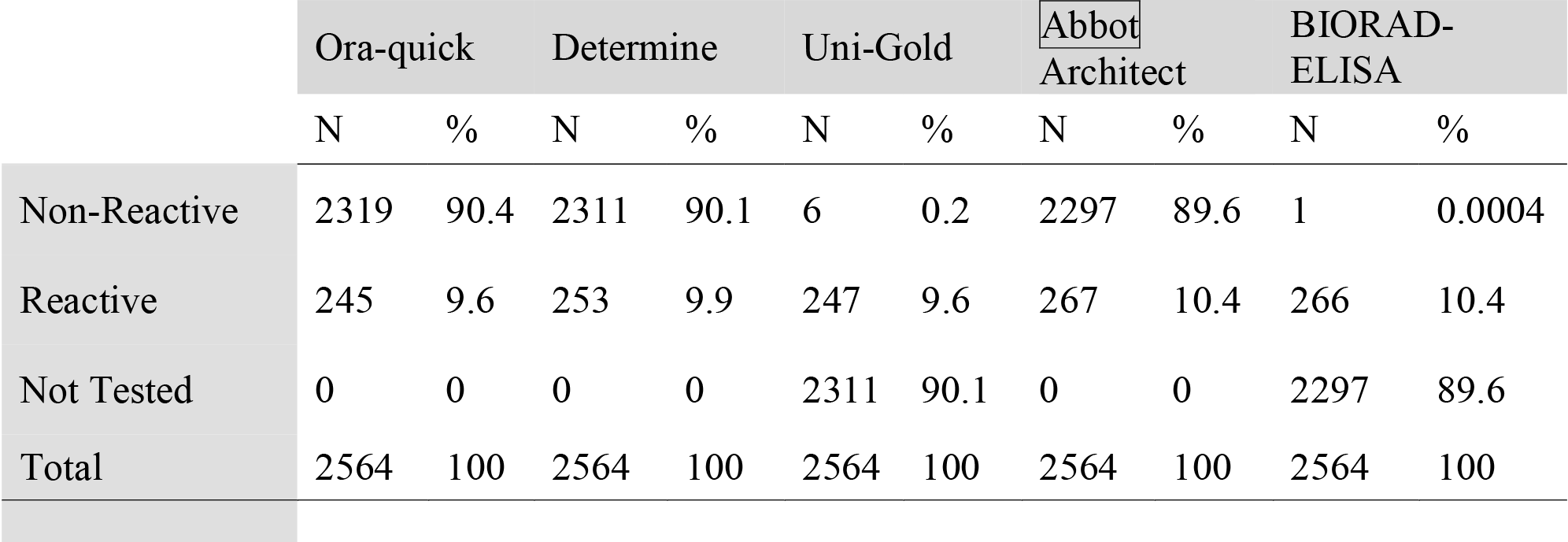
Results of the 5 HIV tests being investigated.

Specimen of participants who were tested on all the 5 HIV tests were included in the analysis. In addition to the 3 Point of Care rapid tests, a blood sample was collected from each participant and sent to the laboratory for testing on Abbot Architect, all positive results on Abbot were confirmed using Genscreen Biorad ELISA. Variables analyzed include baseline demographics and rapid test results. Measurement of each tests’ sensitivity and specificity were compared and reported with 95% confidence intervals. Sensitivity and specificity of Ora-Quick and Determine was measured compared to HIV positive serostatus. HIV positive serostatus was defined by a positive result on the Abbot Architect Assay.

Table 3 shows the results of the comparison of the test results from the 5 HIV tests being investigated

### Ora-Quick

Of the 2564 samples tested on this test, 2319 (90.4 %) were non-Reactive and 245
(9.6%) were Reactive. All samples had a result.

### Determine

all samples tested on Ora-quick were tested on Determine, out of the 2564 samples tested, 2311 (90.1%) were non-Reactive and 253 were Reactive.

### Uni-Gold

Since this was used as a confirmatory test, only samples Reactive on the Determine test were tested on Uni-Gold. 253 samples Reactive on Determine were tested on Uni-gold, 247 were Reactive and 6 samples were non-Reactive. 2311 were not tested because they were Non-Reactive on Determine.

### Abbot Architect

The Abbot Architect was the gold standard test were all above tests were compared to. All 2564 samples were tested on this test, 2297 samples were non-Reactive, and 267 samples were Reactive.

### Genscreen Ultra ELISA

This was a confirmatory test for the Abbot Architect test. Only samples Reactive on Abbot were tested on this test. Of the 267 samples tested, 266 were Reactive and 1 which had a low signal to cut off on Abbot Architect tested Negative. This 1 discordant result was tested on Geenius confirmatory test, and it was Non-Reactive, showing that the Genscreen ultra ELISA is slightly more sensitive than the Abbot Architect test.

The Abbot Architect test was the reference standard, it had 2297 negative results and 267 positive results, when compared to the other Test result (Table 7). When OraQuick results were compared to the gold standard, it was observed that of the 267 reactive results, OraQuick only detected 245 (91.8%) positive results and missed 22 (8.2%). When compared to Abbot Architect results, Determine 249 (93.3%) reactive and 18 tests negative on Determine but positive on ABBOT. The 249 Reactive results on Determine were confirmed on the UniGold test kit, of the 249 tested on UniGold, 247 were reactive and 2 gave false positive results. Compared to Abbot Architect results, UniGold was Reactive on 247 (92.5%) tests and gave 20 (7.5%) non-reactive results.

Table 4 shows results of the HIV tests under investigation compared with the gold standard.

**Table 4:** Comparison of the results of OraQuick, Determine and UniGold to the gold standard.

| Results of 3 Rapid Diagnostic tests compared to Abbot fourth generation test |  |  |  |  |
| --- | --- | --- | --- | --- |
| RAPID DIAGNOSTIC TEST RESULTS |  | Abbot Architect Results |  |  |
|  |  | Non-Reactive | Reactive | Total |
| OraQuick Result | Non-Reactive | 2297 | 22 | 2319 |
|  | Reactive | 0 | 245 | 245 |
|  | Total | 2297 | 267 | 2564 |
| Determine Result | Non-Reactive | 2293 | 18 | 2311 |
|  | Reactive | 4 | 249 | 253 |
|  | Total | 2297 | 267 | 2564 |
| Unigold Result | Non-Reactive | 4 | 2 | 6 |
|  | Reactive | 0 | 247 | 247 |
|  | Not done | 2293 | 18 | 2311 |
|  | Total | 2297 | 267 | 2564 |

### OraQuick Results

the tests that were Non-Reactive on both OraQuick and ABBOT were 2297(89.6%), 245(9.6%) were Reactive on both tests. 22(0.9%) tests were Reactive on ABBOT and non-reactive on OraQuick, meaning that ABBOT picked up 22(0.9%) more positive tests that OraQuick tested as non-reactive. 4(0.2%) tests were Non-reactive on Oraquick but reactive on Determine and Unigold.

#### OraQuick Rapid Test breakdown with reference to ABBOT test results

**Figure 2:**
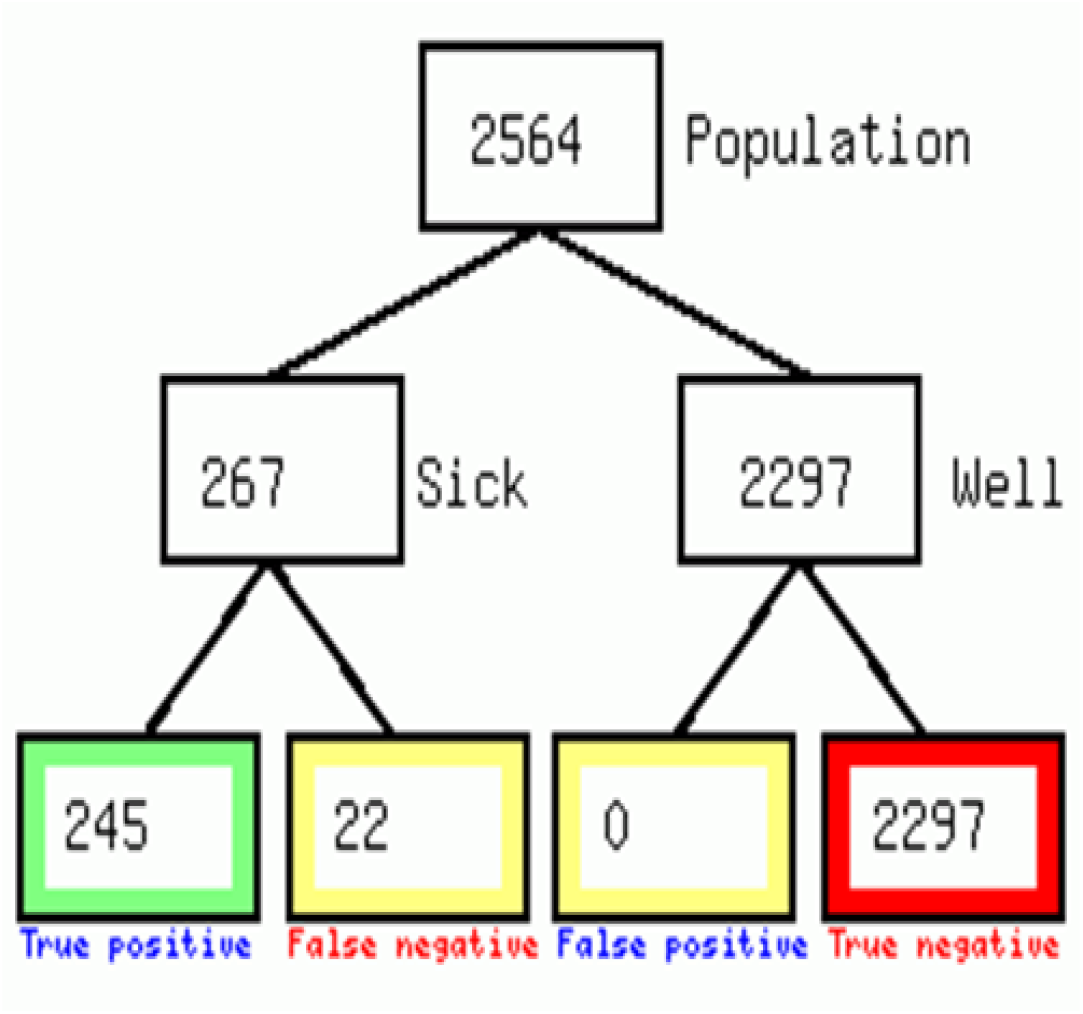
Summary of OraQuick *ADVANCE*® Rapid HIV-1/2 Antibody Test results.

### Determine Results

2293(89.4%) tests were non-reactive on both determine and Abbot Architect. 249 (9.7%) tests were reactive on both Abbot Architect and Determine. 18(0.7%) were Reactive on Abbot Architect and Non-reactive on Determine, showing that Abbot Architect has a higher sensitivity compared to Determine.^17^There were 4 tests that were reactive on Determine and non-reactive on Abbot Architect the reference test, showing a 0.2% low specificity rate of Determine.

#### Determine Rapid Test breakdown with reference to ABBOT test results

**Figure 3:**
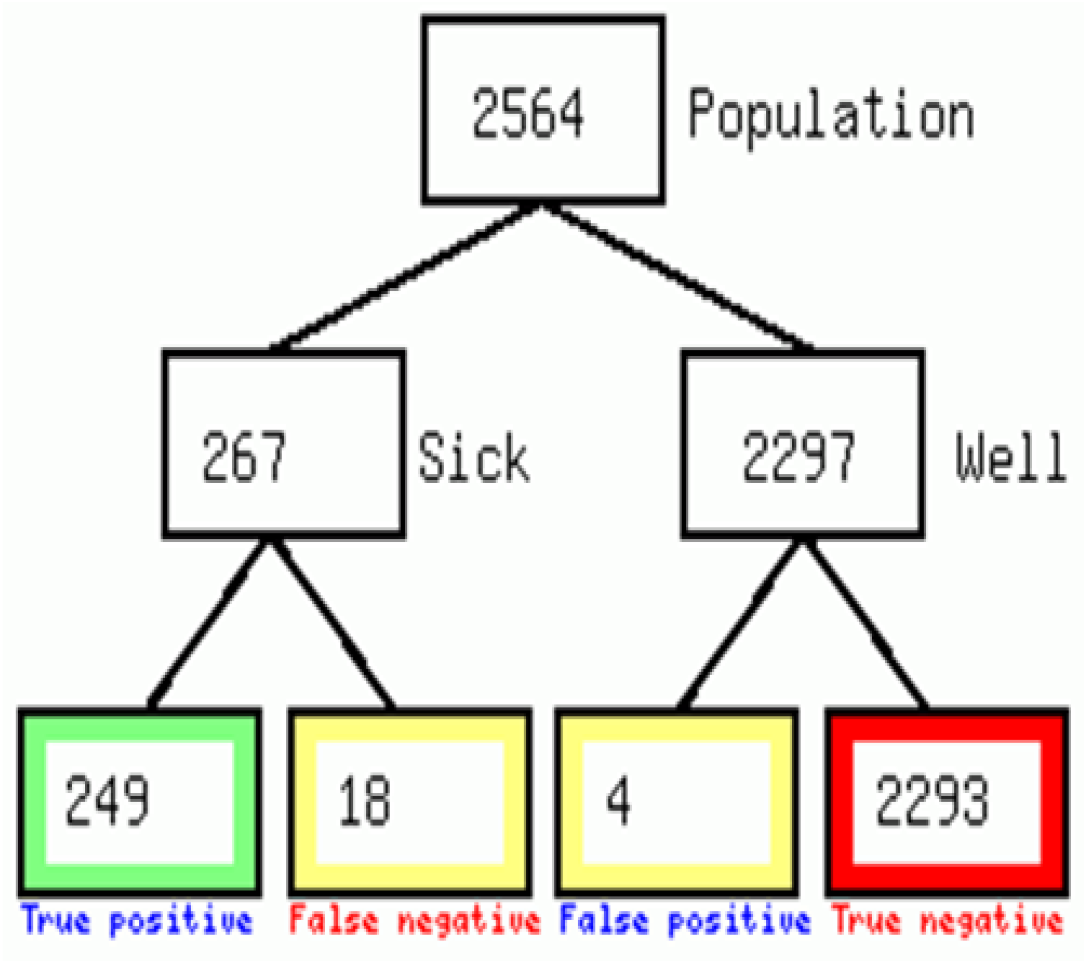
Summary of Abbot Determine™ HIV-1/2 antibody test results.

### UniGold Results

this was used as a confirmatory test for the Determine rapid tests, therefore only the tests reactive on Determine were tested on UniGold. Of the 253(9.9%) tests tested on UniGold, 2(0.9%) were Non-Reactive on Unigold but reactive on Determine and Abbot Architect.

#### Uni-Gold™ Recombigen® HIV-1/2 Test breakdown

**Figure 4:**
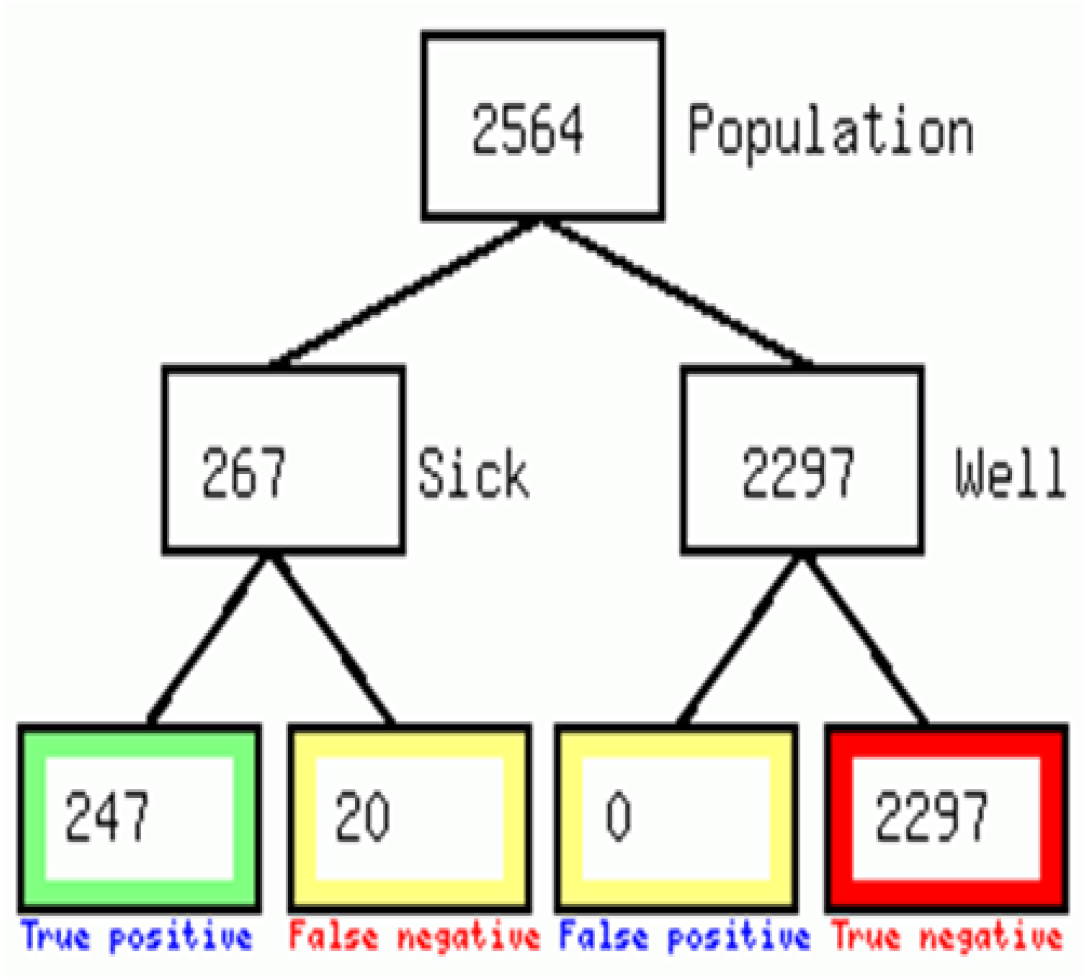
Summary of Uni-Gold™ Recombigen® HIV-1/2 Rapid test results.

When all 3 rapid tests were compared to Abbot Architect, we see that OraQuick gives 22(0.86%) false negatives, UniGold gives 20(0.78%) false negatives and Deterimine had 18(0.70%) false negatives and 4(0.2%) false positives. Of the 3 RDTs, Determine is the most accurate with the highest Sensitivity (93.3%), then UniGold with 92.5% and lastly OraQuick with 91.7% sensitivity, which show a similar result pattern to a study by Kashoshi. Plasma of the 2564 participants was tested on the Abbot Architect analyzer, 2297 (89.6%) were negative and 267 (10.4%) were positive as shown in the table 5. 2,311 (90.5%) had both a negative OraQuick and Determine test result. 245 (91.8%) participants had both a positive Determine and OraQuick test result. The sensitivity and specificity of the OraQuick test were 91.7 (95%CI) and 100 (95%CI), respectively when compared to HIV positive ABBOT test (Table 5). The Sensitivity and Specificity of Determine test were 93.3% and 99.8% respectively.

**Table 5:** Sensitivity, Specificity, Positive predictive value (PPV) and Negative predictive value (NPV) of the 4 Assays.

| Test (Assay) | OraQuick | UniGold | Determine | Abbot Architect |
| --- | --- | --- | --- | --- |
| No of positive results(a) | 245 | 247 | 249 | 267 |
| No of Negative results(d) | 2319 | 2313 | 2311 | 2297 |
| False Negative results© | 22 | 20 | 18 | 0 |
| False Positive results(b) | 0 | 0 | 4 | 0 |
| Sensitivity (%) | 91.8 | 92.5 | 93.3 | 100 |
| Specificity (%) | 100 | 100 | 99.8 | 100 |
| PPV (%) | 100 | 100 | 98.4 | 100 |
| NPV (%) | 99.1 | 99.1 | 99.2 | 100 |

### Discordant results

There was a total of 26(1%) discordant Determine and OraQuick test results. 18 (69.2%) were Determine negative and Abbot Architect positive, 8 (30.8%) were negative on Oraquick and positive on Determine, of the 8, 4 (15.4%) were Determine positive but negative on Abbot Architect and Oraquick and the other 4 (15.4%) were Oraquick negative and positive on both Determine and Abbot. Uni-GoldTM test had less positivity when compared to the total true positive tests. Of the 267 total reactive tests, Unigold detected 247 (92.5%) positives whereas Determine detected 249 (93.3%) positives and Oraquick detected 245 (91.8%).

### Overall Description of the Analyzed Data

Of the 2564 participants who gave a sample, 1521 (59.4%) were female with a mean age of 28.9 years (95% CI: 28.3 – 29.5). 1043 (40.6%) were male with a mean age of 30.9 years (95% CI: – 31.6). 267/2564 samples (10.4%) were found to be HIV-positive by the 4th generation ABBOT Architect HIV Ag/Ab assay. In contrast, only 245/2564 (9.6%), 249/2564 (9.7%), and 247/2564 (9.6%) samples tested positive by OraQuick *ADVANCE*® Rapid HIV-1/2 Antibody Test, Abbot Determine™ HIV-1/2 antibody test and Uni-Gold™ Recombigen® HIV-1/2 respectively (Table 5). When compared to the 4th generation Abbott Architect HIV Ag/Ab Combo assay, there were 22 HIV+ samples that were missed (“false negative”) by all three RDTs under evaluation, but only 4 samples had a reactive result (“false positive”) by Alere Determine HIV-1/2 which were negative on and Uni-Gold HIV which was not confirmed by the 4th generation Abbott Architect HIV Ag/Ab Combo assay.

### Accuracy of the Different Tests

As shown in Table 5, OraQuick *ADVANCE*® Rapid HIV-1/2, Alere Determine HIV-1/2 and Uni-Gold Ultra HIV, at 95% CI had Sensitivities of: 91.8%, 93.3% and 92.5% respectively, of which when compared to the sensitivities reported by the manufacturers is quite low despite manufacturers reports of high sensitivity and specificity for these RDTs.^18^ Generally, the performance of HIV rapid diagnostic testing in a population is influenced by Positive and Negative Predictive values of the RDT kits used. Thus sensitivity and specificity results obtained from test kit evaluation studies to licensing and marketing of the kit will not necessarily be achieved in practice.^19^

The specificities of OraQuick *ADVANCE*® Rapid HIV-1/2 and Uni-Gold HIV were the same (100.0%; 95% CI: 98.8 -100.0) but slightly different from Alere Determine HIV-1/2 (99.8%). Negative predictive values (at 95% CIs) were 99.1, 99.2 and 99.1 for OraQuick *ADVANCE*® Rapid HIV-1/2, Alere Determine HIV-1/2, and Uni-Gold Ultra HIV respectively similar to Kashoshi study findings.^17^

The results shown in table 5 of the reference test-4th generation Abbot Architect HIV Ag/Ab assay demonstrating high sensitivity and maximum positive predictive values. Kappa statistic showed almost perfect agreement between the 4th generation Abbot Architect HIV Ag/Ab and three RDTs: OraQuick *ADVANCE*® Rapid HIV-1/2 (k = 0.952; 95% CI: 0.932 – 0.972), Alere Determine HIV-1/2 (k = 0.953; 95% CI: 0.933 – 0.972), and Uni-Gold Ultra HIV (k = 0.957; 95% CI: 0.938 – 0.976).

## DISCUSSION

This study has established that the three 3rd generation HIV RDTs used at Point of Care for HIV testing though quite sensitive, still fail to detect a significant number of acute infections. About 1 in every 10 people tested using 3^rd^ generation RDTs goes home with false negative results. The identification of persons with acute HIV infection represents a significant challenge, owing to the absence of antibodies in the earliest stages, limitations of standard rapid tests to detect p24 or HIV RNA, and logistical and cost issues with p24 antigen and HIV RNA.^19^ In resource-limited or point-of-care settings, rapid diagnostic tests (RDTs), that aim to simultaneously detect HIV antibodies and p24 capsid (p24CA) antigen with high sensitivity, can pose important alternatives to screen for early infections.^20^

Given the brief window of acute HIV infection, the acute HIV prevalence in a population is very low at any given point in time, even among relatively high-risk groups. Consequently, a test must have exceptional performance characteristics to be useful, especially without additional confirmatory testing.^19^ The Determine®HIV-1/2 Ag/Ab Combo rapid test has been marketed as such a test, because it is relatively inexpensive and can be administered at the point of care.^19^ Although the performance characteristics of the Determine®HIV-1/2 Ag/Ab Combo rapid test were insufficient for use in clinical setting, field evaluation in other settings may be warranted to assess whether the test performs better. The Determine®HIV-1/2 Ag/Ab Combo rapid test has been previously evaluated with stored serum or plasma specimens.^21^

The study is consistent with the findings by Kashosi who found that when compared to the laboratory-based ELISA HIV Ag/Ab assay, the currently used 3rd generation HIV RDTs showed poor field accuracy.^17^ According to their Manufacturers, RDTs have high sensitivity and specificity for licensing purposes and subsequent World Health Organization (WHO) prequalification.^18^ Despite this, HIV RDTs have some limitations since they cannot identify persons who are within the window period of an acute HIV infection because these individuals have not yet developed HIV-specific antibodies.^22, 23^ Such individuals are highly infectious due to concurrent high plasma as well as vaginal and semen HIV-1 viral load.^3, 24^

The study found an unacceptably low sensitivity yet high specificity, and a moderate agreement for all three HIV RDTs evaluated, when compared to the 4th generation Abbot Architect HIV Ag/Ab assay. These results are in line with findings from Ethiopia by Dessie and Bossuyt in which Alere Determine HIV-1/2 showed low sensitivity (60.5%). ^25,26^

This study highlights the important issue of possible false-negative HIV test results, which in part may be explained by the failure of these RDTs to diagnose HIV during the window.^17^ During this period, HIV infection can be detected only by tests detecting also viral antigen such as the laboratory-based 4th generation ELISA.^22^ The baseline HIV prevalence in the population influences the negative and PPV and needs to be considered in the interpretation of HIV RDT results. On the other hand, it is not certain that the currently available 4th generation HIV RDTs would perform better based on recent published data showing that the HIV p24 antigen detection component of some 4th generation RDTs also lacks analytical and diagnostic sensitivity.^26, 27^ According to Kashosi it seemed unlikely that the false-negative samples missed by HIV RDTs in the study, but which were positive for p24 antigen and/or HIV antibodies by the 4th generation ELISA, might represent very early infections (p24 antigen only) or somewhat later ones with antibodies at a low level, not detectable by the RDTs.^17^ Given the qualitative results generated by the laboratory-based 4th generation ELISA test used and the lack of resources for p24 antigen or nucleic acid testing (NAT) in Eastern DRC, Kashosi could not resolve this uncertainty.^17^

Other possible causes that may have caused false-negative results in this study include divergent HIV sub-types, human error-such as the addition of insufficient specimen or too much buffer when procedures are handled by non-trained or unsupervised staff.^28^ transportation or storage of test kits outside of recommended conditions, leading to possible denaturation of reagents or test devices, and the use of expired reagents or test devices.^11^

## CONCLUSION AND RECOMMENDATIONS

This study set out to investigate the performance of HIV Rapid Diagnostic Tests used in Zambia. The results reveal challenges with the third generation RDTs’ ability in detecting acute positive cases. The results suggest that; an alternative HIV testing algorithm, which includes a fourth-generation Determine® HIV-1/2 Ag/Ab Combo rapid test with a better sensitivity and specificity be introduced^19^ Further, there is need to develop and introduce a point-of-care test with adequate sensitivity and specificity for acute HIV detection. Hence the use and scale-up of emerging low cost and point-of-care tests, such as the highly sensitive and easy-to-handle molecular HIV diagnostic tests Xpert HIV-1 Qualitative and Xpert HIV-1 Viral Load Test (e.g., Alere qHIV1/2 detect) should be considered.

## Supporting information

strobe check list

## Data Availability

All data produced in the present study are available upon reasonable request to the authors

## ETHICS APPROVAL AND DISSEMINATION

Ethical approval was granted by the University of Lusaka, medical Ethics Committee, this included a data extraction tool and data management plan to ensure adherence to general data Protection and health research regulations. The end study results will be published in an open access peer-reviewed journal.

## AUTHOR CONTRIBUTIONS

LM

- Work conception. Data acquisition, analysis and interpretation.
- Manuscript drafting.
- Approval of final manuscript.
- Accountable for all aspects of the work regarding its accuracy or integrity.

AT

- Work conception, Data analysis and interpretation.
- Manuscript drafting.
- Approval of final manuscript.
- Accountable for all aspects of the work regarding its accuracy or integrity.

MZ

- Manuscript drafting.
- Critical revisions for intellectual content.
- Data interpretation
- Approval of final manuscript.
- Accountable for all aspects of the work regarding its accuracy or integrity.

PJC

- Manuscript drafting.
- Critical revisions for intellectual content.
- Data interpretation
- Approval of final manuscript.
- Accountable for all aspects of the work regarding its accuracy or integrity.

## ACKNOWLEDGEMENTS

This article is a part of the master’s thesis submitted to the University of Lusaka in partial fulfillment of the requirements for the degree of Master of Public Health. Special acknowledgements to ZAMBART for the opportunity to use the STAR data for this study.

## REFERENCES

1. Piot, P. Quinn, T. C. (2013) Response to the AIDS Pandemic - A Global Health Model, New England Journal of Medicine June 6, 2013 368(23):2210. https://www.nejm.org/doi/full/10.1056/NEJMra1201533.

2. World Health Organization (2018) Consolidated Guidelines on HIV Testing Services Available at: http://apps.who.int/iris/bitstream/10665/ Accessed: 28 April 2021.

3. Cohen S.M, Cynthia L. Gay, Michael P. Busch, Frederick M. Hecht. (2010) The Detection of Acute HIV Infection, The Journal of Infectious Diseases, Volume 202, Pages S270–S277, https://doi.org/10.1086/655651

4. World Health Organization. (2014) HIV/AIDS. Global situation and trends. Available at http://www.who.int/gho/hiv/en/ accessed on 13th March 2021.

5. Patel, P., Bennett, B., Sullivan, T., Parker, M. M., Heffelfinger, J. D., Sullivan, P. S., & CDC AHI Study Group (2012). Rapid HIV screening: missed opportunities for HIV diagnosis and prevention. Journal of clinical virology: the official publication of the Pan American Society for Clinical Virology, 54(1), 42–47. https://doi.org/10.1016/j.jcv.2012.01.022

6. Daskalakis D. (2011). HIV diagnostic testing: evolving technology and testing strategies. Topics in antiviral medicine, 19(1), 18–22.

7. Mitchell .E. O, Greg Stewart, Olivier Bajzik, Mathieu Ferret, Christopher Bentsen, M. Kathleen Shriver. (2013). Performance comparison of the 4th generation Bio-Rad Laboratories GS HIV Combo Ag/Ab EIA on the EVOLIS™ automated system versus Abbott ARCHITECT HIV Ag/Ab Combo, Ortho Anti-HIV 1+2 EIA on Vitros ECi and Siemens HIV-1/O/2 enhanced on Advia Centaur, Journal of Clinical Virology,Volume 58, Supplement 1, Pages e79–e84, https://doi.org/10.1016/j.jcv.2013.08.009.

8. Bentsen C, McLaughlin L, Mitchell E, Ferrera C, Liska S, Myers R, Peel S, Swenson P, Gadelle S, Shriver MK. (2011). Performance evaluation of the Bio-Rad Laboratories GS HIV Combo Ag/Ab EIA, a 4th generation HIV assay for the simultaneous detection of HIV p24 antigen and antibodies to HIV-1 (groups M and O) and HIV-2 in human serum or plasma. J Clin Virol 52 Suppl 1: 57–61.

9. Chavez P, Wesolowski L, Patel P, Delaney K, Owen SM. (2011). Evaluation of the performance of the Abbott ARCHITECT HIV Ag/Ab combo assay. J Clin Virol 52 Suppl 1: 51–55.

10. Alere Limited. (2014). Alere Determine® HIV-1/2; 20DetermineW%20HIV%20eng.pdf, Alere Determine HIV 1/2 Ag/Ab Combo, package insert. Waltham, MA, USA; 2014.

11. Lewis, J. M., Macpherson, P., Adams, E. R., Ochodo, E., Sands, A., & Taegtmeyer, M. (2015). Field accuracy of fourth-generation rapid diagnostic tests for acute HIV-1: a systematic review. AIDS (London, England), 29(18), 2465–2471. https://doi.org/10.1097/QAD.0000000000000855

12. Centers for Disesase Control and Prevention. (2018). Rapid HIV tests suitable for use in nonclinical settings (requires CLIA waiver certificate). Centers for Disesase Control and Prevention. http://www.cdc.gov/hiv/pdf/testing_nonclinical_clia-waived-tests.pdf [Accessed 27 April 2021].

13. Masciotra S, McDougal JS, Feldman J, et al. Evaluation of an alternative HIV diagnostic algorithm using specimens from seroconversion panels and persons with established HIV infections, J Clin Virol, 2011, vol. 52 uppl 1(pg. S17–22)

14. Zambia Demographic and Health Survey. (2018). HIV/AIDS Related Knowledge, Attitudes and Behaviour, Rockville, Maryland, USA: Zambia Statistics Agency, Ministry of Health, University Teaching Hospital Virology Laboratory and ICF International, pg 223–257.

15. Ayles H., Bond V., Chintu. N., Handima N., Kapaku K., Maluzi K., et al. (2016). HIV Self Testing Africa-Zambia. A clinical performance study of self-testing using self-collected oral fluid transudate and the OraQuick® HIV Self-Test.

16. Boadu, R., Darko, G., Nortey, P., Akweongo P., Sarfo B., (2016). Assessing the sensitivity and specificity of First Response HIV-1-2 test kit with whole blood and serum samples: a cross-sectional study. AIDS Res Ther 13, 9 https://doi.org/10.1186/s12981-016-0092-0

17. Kashosi T. M, et al. (2018), The journal of infection in developing countries, Field accuracy of HIV rapid diagnostic tests for blood donor screening, Bukavu, Eastern Democratic Republic of the Congo.

18. Piwowar-Manning, E. M., Tustin, N. B., Sikateyo, P., Kamwendo, D., Chipungu, C., Maharaj, R., Mushanyu, et al. (2010). Validation of Rapid HIV Antibody Tests in 5 African Countries. Journal of the International Association of Physicians in AIDS Care, 9(3), 170–172. https://doi.org/10.1177/1545109710368151

19. Rosenberg, N. E., Kamanga, G., Phiri, S., Nsona, D., Pettifor, A., Rutstein, S. E., Kamwendo, D., Hoffman, I. F et al. (2012). Detection of acute HIV infection: a field evaluation of the determine® HIV-1/2 Ag/Ab combo test. The Journal of infectious diseases, 205(4), 528–534. https://doi.org/10.1093/infdis/jir789

20. Wratil, P. R., Rabenau HF, Eberle J. (2020). Comparative multi-assay evaluation of Determine™ HIV-1/2 Ag/Ab Combo rapid diagnostic tests in acute and chronic HIV infection. Medical microbiology and immunology, 209(2), 139–150. https://doi.org/10.1007/s00430-019-00655-0).

21. Peeling RW, Mabey D. Point-of-care tests for diagnosing infections in the developing world, Clin Microbiol Infect, 2010, vol. 16 (pg. 1062–9)

22. Patel P, Mackellar D, Simmons P. (2010). Detecting Acute Human Immunodeficiency Virus Infection Using 3 Different Screening Immunoassays and Nucleic Acid Amplification Testing for Human Immunodeficiency Virus RNA, 2006-2008. Arch Intern Med. 170(1):66–74. doi:10.1001/archinternmed.2009.445

23. Zetola N.M., Pilcher D.C. (2007). Diagnosis and Management of Acute HIV Infection, Infectious Disease Clinics of North America,Volume 21, Issue 1, Pages 19–48, ISSN 0891-5520, https://doi.org/10.1016/j.idc.2007.01.008. (2007). Diagnosis and management of acute HIV infection. Infect Dis Clin North Am 21: 19-48.

24. Morrison CS, Demers K, Kwok C, et al. Plasma and cervical viral loads among Ugandan and Zimbabwean women during acute and early HIV-1 infection, AIDS, 2010, vol. 24 (pg. 573–82)

25. Bossuyt PM, Reitsma JB, Bruns DE, Gatsonis CA, Glasziou PP, Irwig LM, (2003). Towards complete and accurate reporting of studies of diagnostic accuracy: The STARD Initiative. Ann Intern Med 2003;138:40-4.: the STARD initiative. BMJ 326: 41–44.

26. Dessie, A, Abera, B, Walle, F, Wolday, D, Tamene, W. (2008). Evaluation of determine HIV-1/2 rapid diagnostic test by 4th generation ELISA using blood donors’ serum at Felege Hiwot referral Hospital, Northwest Ethiopia. Ethiop Med J; 46: 1–5.

27. Taegtmeyer, M, MacPherson, P, Jones, K. (2011). Programmatic evaluation of a combined antigen and antibody test for rapid HIV diagnosis in a community and sexual health clinic screening programme. PLoS One 2011; 6(11): e28019–e28019.

28. Beelaert G, Fransen K. (2010). Evaluation of a rapid and simple fourth-generation HIV screening assay for qualitative detection of HIV p24 antigen and/or antibodies to HIV-1 and HIV-2. J Virol Methods 168: 218–222.

29. Michaeli M, Wax M, Gozlan Y, Rakovsky A, Mendelson E, Mor O. Evaluation of xpert HIV-1 qual assay for resolution of HIV-1 infection in samples with negative or indeterminate geenius HIV-1/2 results. J Clin Virol. 2016; 76: 1–3.

30. Moyo S, Mohammed T, Wirth KE, Prague M, Bennett K, Holme MP, et al. (2016). Point-of-care Cepheid Xpert HIV-1 viral load test in rural African communities is feasible and reliable. J Clin Microbiol 54:3050–3055.

